# Independent, Diplotype and Haplotype Association Analyses of the Selected *MTHFR* SNPs with the Risk of Breast Cancers in a South-Asian Population

**DOI:** 10.1101/2021.03.13.21253512

**Authors:** Sadia Ajaz, Sani-e-Zehra Zaidi, Saleema Mehboob Ali, Aisha Siddiqa, Muhammad Ali Memon, Aiysha Abid, Shagufta Khaliq

## Abstract

**PURPOSE:** MTHFR is a pivotal enzyme in the folic acid cycle. In specific populations, two functional SNPs in the encoding gene, i.e. rs1801133 (677C/T) and rs1801131 (1298A/C), have shown associations with different diseases including cancers. In the present study, the role of these SNPs was analyzed in breast cancer cases from Pakistani population.

**PARTICIPANTS AND METHODS:** The pilot study includes 187 participants with 124 breast cancer patients and 63 matched individuals as controls. PCR-RFLP methods validated by Sanger sequencing were used for the polymorphic investigations. Hardy-Weinberg Equilibrium was tested by chi-squared goodness-of-fit test. Independent and combined associations were evaluated by Cochran-Armitage trend-test. Semi-parameteric haplotype analysis was carried out and odds ratios with 95% confidence interval were tabulated. Linkage disequilibrium between two loci was calculated. p-values <0.05 were significant.

**RESULTS:** Both *MTHFR* 677C/T and *MTHFR* 1298A/C SNPs were in Hardy-Weinberg Disequilibium. Cochran-Armitage trend test showed lack of independent associations of these SNPs with the risk for breast cancers. The diplotype analysis revealed that 677CC+1298AC increased the risk of breast cancers significantly [OR: 2.553 (95% CI: 1.177-5.541)], while 677CT+1298AA had a protective effect [OR: 0.537 (95% CI: 0.404-0.713)]. Haplotype analysis did not show any significant association. Interestingly, despite the proximity, these loci were not linked (r^2^ = 0.042 and 0.046 in cases and controls, respectively).

**CONCLUSION:** Here, we report association analysis of two putative candidate SNP markers in the *MTHFR* gene with breast cancers in a South-Asian population. To the best of our knowledge, two diplotype combinations show unique associations with breast cancer susceptibility in this population, which have not been reported earlier. The study implies translational potentials of these polymorphisms for breast cancer management.

## INTRODUCTION

Breast cancers have high rates of morbidity and mortality worldwide. In the developing countries, breast cancer is the leading cause of cancer-related deaths among females [1]. Pakistan is located in South-Asian region. In this country, the age-specific incidence and mortality rates of breast cancer(s) are among the highest in Asian nations and globally, respectively [2].

One of the major aims of molecular genetics in breast cancers is to identify and quantitate the contribution of disease-associated loci. These loci are likely to encode functional biological products, which contribute in maintaining normal physiology within tissues and organs.

Folate pathway provides protection against neoplastic transformation and progression [3,4]. Malfunction in this pathway leads to genomic instability, aberrant DNA methylation, and the accumulation of toxic carcinogens. Such changes underpin cancer etiology and progression [5-7].

<u>M</u>ethylene <u>T</u>etrahydrofolate <u>R</u>eductase (MTHFR) (UniProtKB-P42898) is a 77kDa key enzyme of the folate pathway. It converts 5-10 methylene tetrahydrofolate (5,10 MTHF) to 5-methyl tetrahydrofolate (5 MTHF) [8]. The latter serves as donor of methylation group in the homocysteine-based re-methylation of methionine. MTHFR concentration occupies a critical position in directing the folate pool towards either homocysteine remethylation or DNA and RNA biosynthesis [9].

Decreased MTHFR activity has been associated with gastrointestinal stromal tumour, neural tube defects, folate sensitivity, MTHFR-deficiency, schizophrenia, as well as dosage and toxicity response to adriamycin and cyclophosphamide [10,11]. The gene for MTHFR is located on chromosome 1p36.3. Among the variants, two functional <u>S</u>ingle <u>N</u>ucleotide <u>P</u>olymorphisms (SNPs), 677C/T (rs1801133) and 1298A/C (rs1801131) encode for thermolabile isoforms of the MTHFR. These variants affect its enzymatic activity. The function is reduced by almost 30% for the *MTHFR* 677C/T variation, while there is almost 60% decrease due to *MTHFR* 1298A/C polymorphism [12]. The 677T-variant results in the substitution of alanine with valine (Ala222Val). In 1298-C variant, glutamic acid is replaced with alanine (Glu429Ala) [13].

In cancer cell, thermolabile isoform of the MTHFR leads to the mis-incorporation of uracil instead of thymine with consequent DNA damage [14]. Additionally, reduced enzymatic activity affects DNA methylation and hence dysregulates the gene expression [15].

The SNPs are located within a few kilo-bases of each other and due to the short distance are expected to be in high degree of linkage disequilibrium (LD) [16, 17]. However, several discrepancies in other closely-related genomic regions have been reported [18]. Therefore, the quantitative value of LD among proximate genetic variations need to be assessed separately for each population. This is essential to map the population-specific contribution of closely situated polymorphisms in the disease phenotype.

Data regarding the role of these functional SNPs in the breast cancers remains inconclusive in different populations [19, 20]. Given this rationale, the present study is a population-centered systematic attempt to investigate the role of these functional *MTHFR* SNPs in breast cancers in Pakistani population.

## METHODOLOGY

### Ethics Statement

The study was conducted in accordance with the Declaration of Helsinki [21]. The project was approved by the ethical review committees (ERCs) of the participating institutions: the independent ERC, International Center for Chemical and Biological Sciences (ICCBS), University of Karachi, Karachi, Pakistan [ICCBS/IEC-016-BS/HT-2016/Protocol/1.0], and the Atomic Energy Medical Centre (AEMC), Jinnah Postgraduate Medical Centre (JPMC), Karachi, Pakistan [Admin-3(257)/2016]. Written informed consents were volunteered by each participant. The ethical approval from controls has been submitted and published earlier [22].

### Study Participants

The pilot study comprised case-control design and included 187 participants. The cases included 124 diagnosed and histologically confirmed primary breast cancer patients. The patients were either first-time visitors or under regular treatment at AEMC, JPMC, Karachi, Pakistan. The project duration was from July 2016 – July 2017. The only exclusion criterion for patients was lack of confirmed biopsy report for breast cancer. In controls, 63 samples of medically-confirmed, healthy individuals were included [22]. The control samples were matched on the basis of age, gender and ethnicity. The exclusion criteria for controls were: age or gender based mismatch, previous diagnosis of any cancer or co-morbidity with any other chronic disease. The participants belonged to Southern-Pakistan.

#### 1.1 Breast Cancer Data

At the time of sampling, participants’ relevant information was obtained through a questionnaire. Data regarding age, ethnicity, place of residence, contact number, family history of cancers, age at menarche, obstetrics and gynaecology history, and if applicable, the age at menopause were collected. Clinical characteristics including, tumor histology, size, grade, stage, axillary lymph node metastasis, ER status, PR status, and Her-2 status were collected from the patients’ medical records. The collection of clinical data depended upon the availability of such information in patients’ medical record file.

### DNA Extraction

DNA samples were extracted using standard phenol-chloroform method with slight modifications [23].

### PCR-RFLP for MTHFR 677C/T

In case of 677C/T variant, the DNA fragment of 198bp was amplified in 25µl total volume. PCR mix contained 1X (NH_4_)_2_SO_4_ PCR buffer, 0.4mM MgCl_2_, 0.2mM dNTPs, 0.4U Taq polymerase, 0.35µM of each primer (Forward primer: 5’-TGAAGGAGAAGGTGTCTGCGGGA-3’; Reverse primer: 5’-AGGACGGTGCGGTGAGAGTG-3’) and 130ng of DNA template in a final reaction volume of 25 µl. Amplification was carried out at annealing temperature (T_a_) of 64°C. Cycling conditions have already been published [22]. 20µl of the amplified product was digested overnight with 10U of *HinfI* and 3.5µl of the recommended buffer (Thermo Scientific^®^, USA). The digested products were run on 10% polyacrylamide gel, stained with ethidium bromide, and observed under UV. Representative gel for genotyping of the *MTHFR* 677C/T is shown in Figure 1.

**Figure 1.**
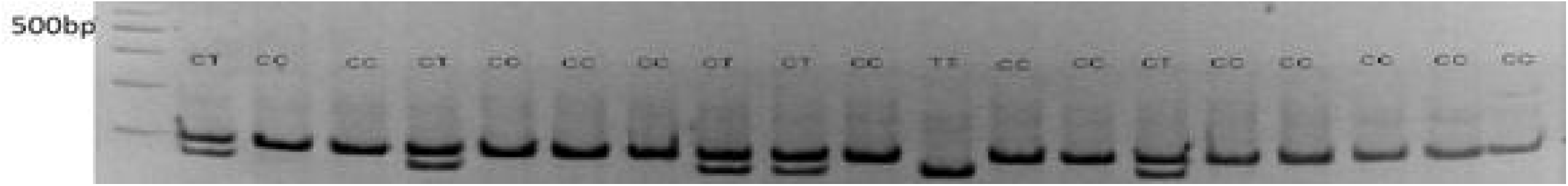
*MTHFR* 677C/T genotyping after digestion with *HinfI* restriction enzyme using 10% polyacrylamide gel stained with ethidium bromide, CC genotype: 198-bp fragment, TT genotype: 175-bp fragment, CT genotype: 198-bp and 175-bp fragments.

### PCR-RFLP for MTHFR 1298A/C

In case of 1298A/C, the DNA fragment of 168bp was amplified. PCR mix of 25µl contained 1X (NH_4_)_2_SO_4_ based PCR buffer with 0.8mM of MgCl_2_, 0.25mM of dNTPs, 2U of Taq polymerase, 1.25µM of each primer (Forward primer: 5’-CTTTGGGGAGCTGAAGGACTACTA-3’; 5’-CACTTTGTGACCATTCCGGTTTG-3’) and 70ng of DNA template. Amplification was carried out at T_a_ of 60°C. Cycling conditions were the same as for 677C/T. 20µl of the amplified product was digested overnight with 2.5U of *MboII* and 2µl of the recommended buffer (Thermo Scientific^®^, USA). The digested products were run on 10% polyacrylamide gel, stained with ethidium bromide, and observed under UV. Representative gel for 1298A/C is shown in Figure 3.

### Validation by Sanger Sequencing

Results of both polymorphisms were validated by Sanger sequencing, commercially. The validation of each methodology by Sanger sequencing for *MTHFR* 677C/T and *MTHFR* 1298A/C is shown in figures 2 and 4 respectively.

**Figure 2.**
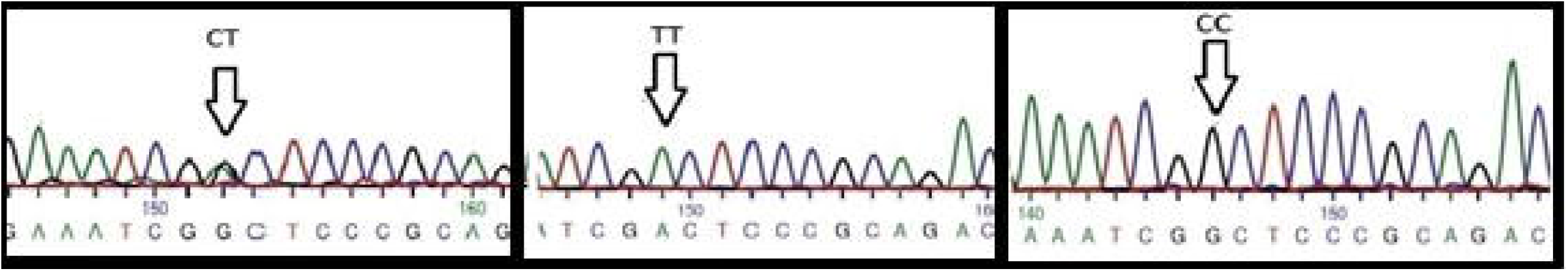
Validation of *MTFR* 677C/T genotyping by Sanger sequencing.

**Figure 1.**
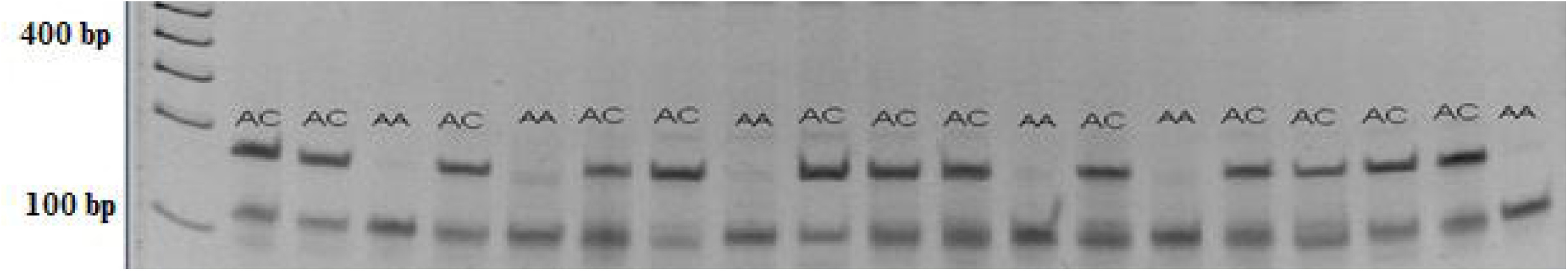
*MTHFR* 1298A/C genotyping after digestion with *MboII* restriction enzyme using 10% polyacrylamide gel stained with ethidium bromide, AA genotype: 56-bp fragment, CC genotype 87-bp fragment, AC genotype: 56-bp and 87-bp fragments.

**Figure 4.**
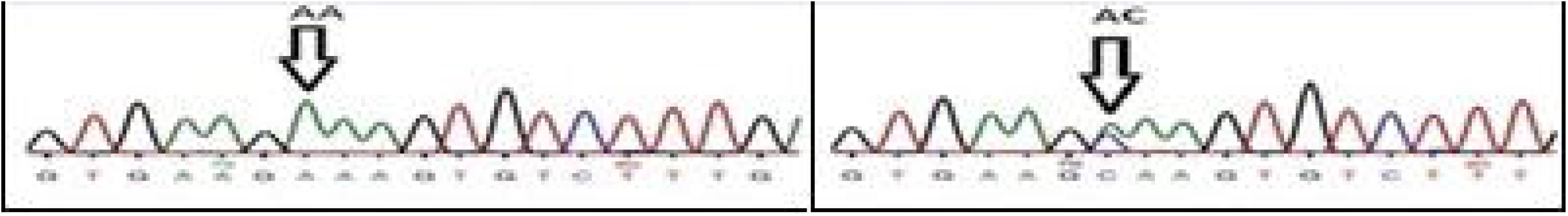
Validation of *MTFR* 1298A/C genotyping by Sanger sequencing.

### Statistical Analyses

Statistical analyses were carried out using IBM (SPSS^®^) v.21.0 software [24]. The genotype and allele frequencies were determined by gene counting method along with the weighted percentages. Genotype distributions among the cases and controls were analyzed for Hardy-Weinberg equilibrium using chi-squared goodness of fit test. The association between qualitative variables such as clinico-pathological characteristics including tumour stages and grade were also assessed by Pearson’s Chi-squared test. Cochran-Armitage trend test was used for the assessment of the breast cancer risk association as Hardy-Weinberg disequilibrium was observed [16]. In order to measure the allelic, genotypic and haplotype risks for breast cancers, odds ratios (OR) with 95% confidence interval (95% CI) were calculated using logistic regression, while association was evaluated by Cochran-Armitage trend test. For all the analyses p-value <0.05 were considered to be significant. The associations were found to be independent of the genetic models.

#### 1.2 Haplotype Estimation and LD Statistics

Estimation of haplotype frequencies and LD analysis were carried out using cubeX webtool [25]. The maximum likelihood estimations were carried out for the calculations of haplotype frequencies. For the estimation of LD between two loci, this programme was used for the calculation of Lewontin’s standardized disequilibrium coefficient (D’), correlation co-efficient (r^2^) and χ^2^ test (with significant value at p<0.05) were used. The results were compared with the Phase 3 (version 5) 1000 Genomes Project data for different populations on LDlink, a National Cancer Institute website tool [26].

## 3 RESULTS

### Participants’ Demographics and Patients’ Clinico-pathological Data

The average age of cases was 44.35 ± 0.932 years while that of the controls was 46.04 ± 0.80 years, respectively. The age difference between two groups was statistically non-signficant (Mann-Whitney test, p-value >0.05). Clinico-pathological data for the studied cohort is shown in Table 1. In summary, majority of the patients had invasive ductal carcinoma (79%) with tumour size >5cm (48%), stage III (55%), and grade 3 (52%) tumours.

**Table 1.** Clinico-pathologic Characteristics of the Breast Cancer Samples.

| *Sr. No.* | *Tumour characteristic* | *Value/ numbers* |
| --- | --- | --- |
| 1. | Tumour Size | <2cm : 11 (9%) |
|  |  | 2-5cm : 52 (43%) |
|  |  | >5cm : 58 (48%) |
| 2. | Tumour Stage | T<sub>1</sub>: 03 (03.84%) |
|  |  | T<sub>2</sub>: 26 (33.33%) |
|  |  | T<sub>3</sub>: 43 (55.12%) |
|  |  | **T<sub>4</sub>:** 06 (07.69%) |
| **3.** | **Tumour Grade** | **G1:** 01 (0.94%) |
|  |  | **G2:** 48 (45.28%) |
|  |  | **G3:** 55 (51.88%) |
|  |  | **G4:** 2 (1.88%) |
| **4.** | **Histopathology** | **IDC:** 88 (79.3%) |
|  |  | **DCIS:** 01 (0.9%) |
|  |  | **LCIS:** 0 |
|  |  | **ILC:** 02 (1.80%) |
|  |  | **DCIS + IDC:** 15 (13.5%) |
|  |  | **Others:** 05 (4.50%) |

IDC: Invasive Ductal Carcinoma; DCIS: Ductal Carcinoma Insitu; LCIS: Lobular Carcinoma In situ; ILC: Invasive Lobular Carcinoma

### Hardy-Weinberg Equilibrium Estimation

The distributions of genotypes and allele frequencies of the *MTHFR* 677C/T and 1298A/C polymorphisms are given in Tables 2 and 3, respectively. χ^2^ values showed that the distributions of genotypes occurred in Hardy-Weinberg disequilibrium for both polymorphisms [*MTHFR* 677C/T: χ^2^ value=3; p-value <0.05(controls) and χ^2^ value=3.39; p-value <0.05(cases)]; [*MTHFR* 1298A/C: χ^2^ value=5.62; p-value <0.05(controls) and χ^2^ value=36.21; p-value <0.05(cases)].

**Table 2.** Distribution of the *MTHFR* 677C/T genotypes and the allele frequencies (with standard errors) in controls and cases (breast cancer patients). Chi-square test, odds-ratios (95% Confidence Interval), and Cochran-Armitage trend test for the risk of breast cancers.

| ***MTHFR***<br>**677C/T**<br>**Polymorphism** | **Controls**<br>**(n = 62<sup>s</sup>)** | **Breast**<br>**cancer**<br>**patients**<br>**(n = 124)** | **Chi-square**<br>**(p-value)** | **Odds-ratios (OR)**<br>**(95% Confidence**<br>**Interval)** | **Cochran-**<br>**Armitage**<br>**Test**<br>**IzI value**<br>**(p-value)** |
| --- | --- | --- | --- | --- | --- |
| **Genotypes** | N (%) | N (%) |  |  | 0.019<br>(0.722) |
| CC | 47 (75.8%) | 90 (72.6%) | Ref. | Ref. |  |
| CT | 12 (19,3%) | 28 (22.6%) | 0.26 (0.70) | 1.22 (0.57 – 2.61) |  |
| **TT** | 03 (4.8%) | 06 (4,8%) | 0.004 (1.00) | 1.04 (0.25– 4.36) |  |
| **Allele Frequencies** |  |  |  |  |  |
| p[C] | 0.85±0.032 | 0.84±0.032 | 0.163 (0.76) | 1.132 (0.62 – 2.07) | 0.022<br>(0.702) |
| q[T] | 0.15±0.032 | 0.16±0.032 |  |  |  |

<sup>§</sup>No result was obtained for 01 control samples.

**Table 3.** Distribution of the *MTHFR* 1298A/C genotypes and the allele frequencies (with standard errors) in controls and cases (breast cancer patients). Chi-square test, odds-ratios (95% Confidence Interval), and Cochran-Armitage trend test for the risk of breast cancers.

| ***MTHFR***<br>**1298 A/C**<br>**Polymorphism** | **Controls**<br>**(n = 63)<sup>§</sup>** | **Breast**<br>**cancer**<br>**patients**<br>**(n = 124)** | **Chi-square**<br>**(p-value)** | **Odds-ratios (OR)**<br>**(95% Confidence**<br>**Interval)** | **Cochran-**<br>**Armitage**<br>**Test**<br>**IzI value**<br>**(p-value)** |
| --- | --- | --- | --- | --- | --- |
| **Genotypes** | N (%) | N (%) |  |  | 0.027<br>(0.818) |
| AA | 21 (33.3%) | 37 (29.8%) | Ref. | Ref. |  |
| AC | 33 (52.4%) | 87 (70.2%) | 1.40 (0.29) | 1.49 (0.77 – 2.92) |  |
| CC | 04 (06.3%) | 00 (0%) | 2.38 (0.14) | 0.32 (0.07– 1.44) |  |

| Allele Frequencies |  |  |  |  |  |
| --- | --- | --- | --- | --- | --- |
| p[A] | 0.85±0.032 | 0.84±0.032 | 0.002 (1.00) | 0.998 (0.62 – 1.57) | 0.029<br>(0.973) |
| q[C] | 0.15±0.032 | 0.16±0.032 |  |  |  |

<sup>§</sup>No results were obtained for 05 control samples.

### Insignificant Associations of *MTHFR* 677C/T and *MTHFR* 1298A/C Polymorphisms with Risk for Breast Cancers

No overall significant associations were found between the polymorphisms and the risk for breast cancers. Statistical analysis of these SNPs with breast cancers are shown in Tables 2 and 3.

### Unique Significant Associations of 677CC*/*1298AC and 677CT/1298AA Diplotypes with Breast Cancer Susceptibility

The nine possible genotype combinations for two *MTHFR* polymorphisms are shown in Table 4. Case-control association analysis was carried out for each combination. The diplotype analysis revealed that 677CC+1298AC increased the risk of breast cancers significantly [OR: 2.553 (95% CI: 1.177-5.541)], while 677CT+1298AA had a protective effect [OR: 0.537 (95% CI: 0.404-0.713)] (Table 4).

**Table 4.**
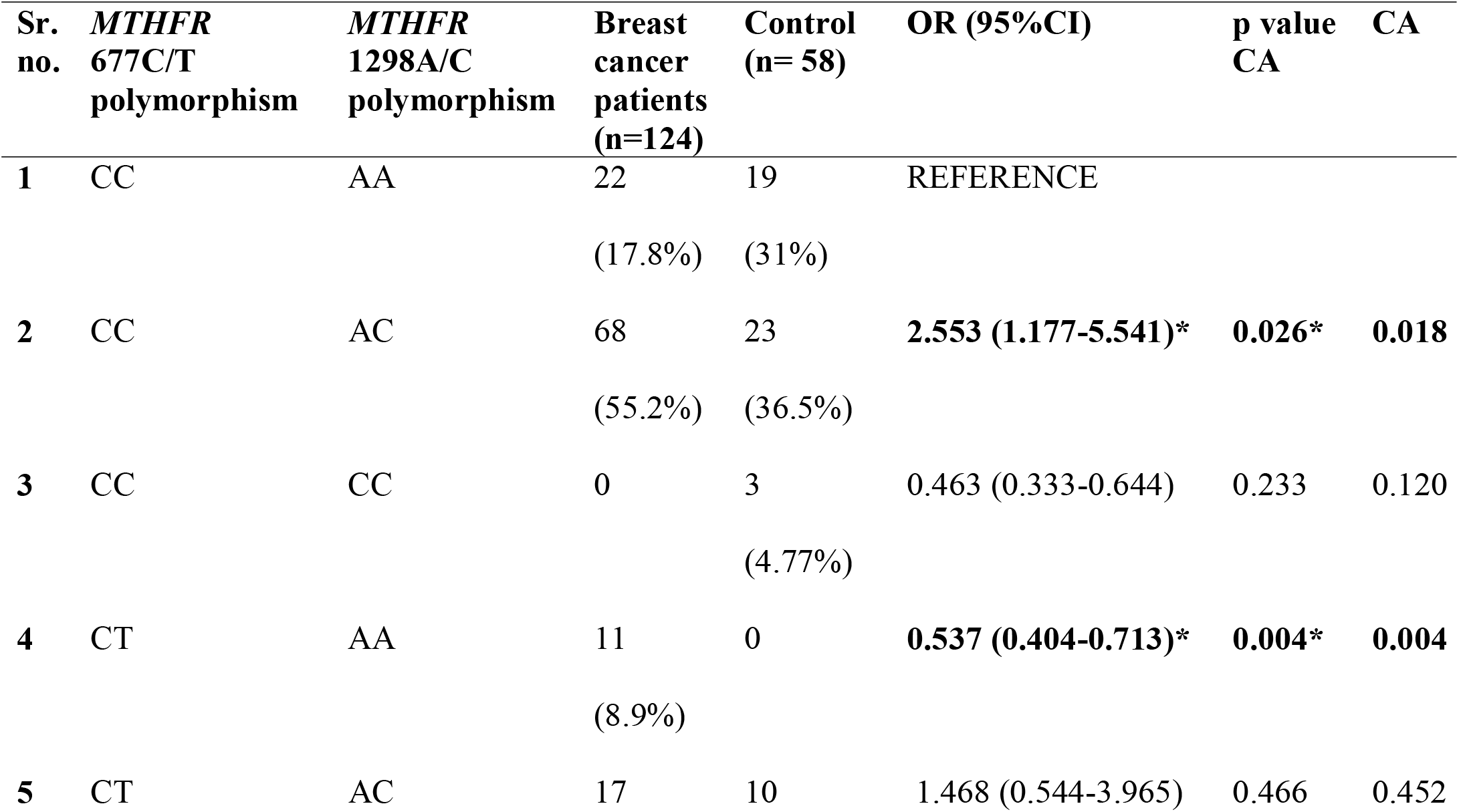
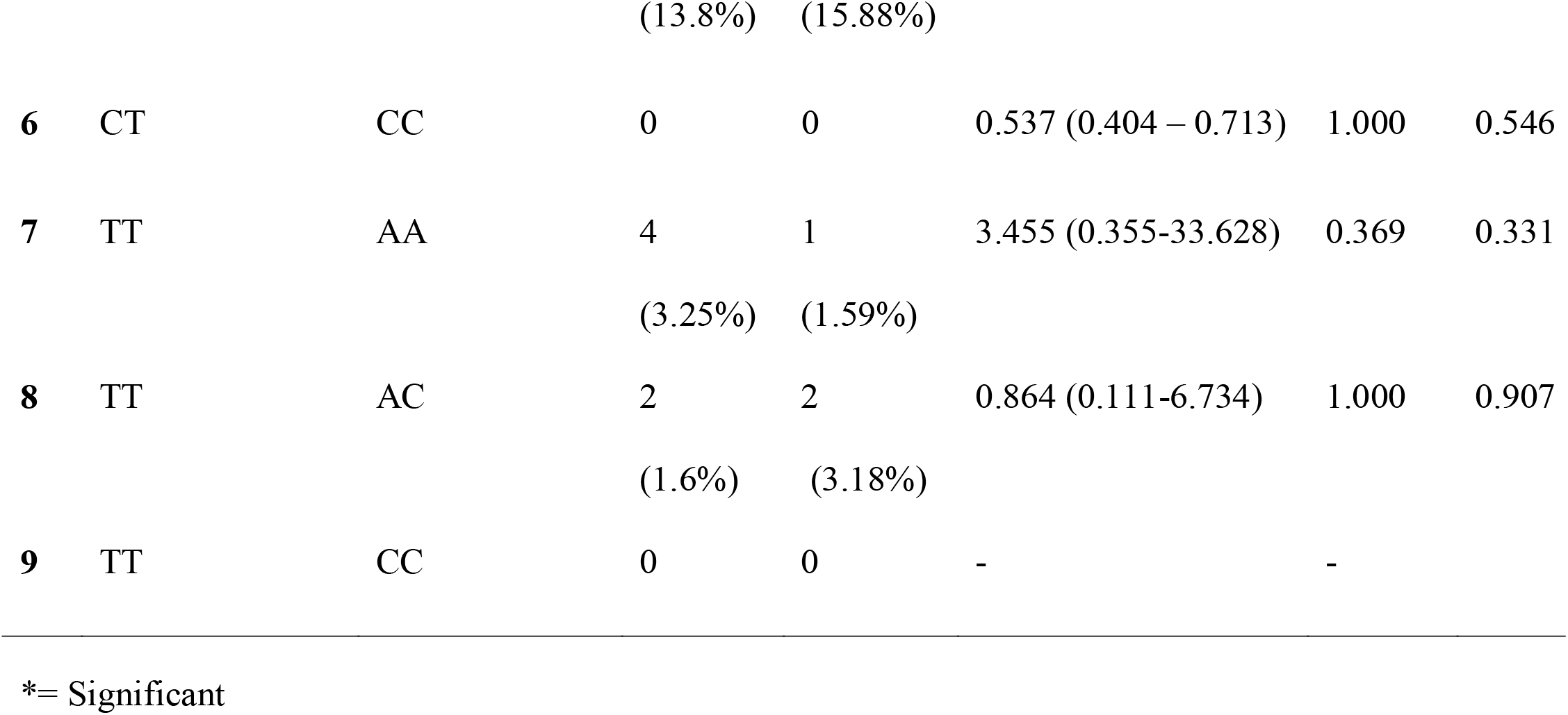
Distribution of combined genotypes of *MTHFR* 677CT and 1298AC polymorphism in control and cases (breast cancer patients) and their risk assessment using OR (95%CI)

### Haplotype and LD Analysis at *MTHFR* 677C/T and 1298A/C Locus

The disequilibrium spread in pairwise allelic combinations at the *MTHFR* 677C/T and 1298A/C loci was quantified by maximum likelihood calculation from the frequency of diploid genotypes. Haplotype frequencies and the LD statistics, D’, r^2^, and χ^2^ values demonstrate that the variants at two loci are not associated (Table 5). None of the haplotypes showed any significant association with breast cancer susceptibility

**Table 5.**
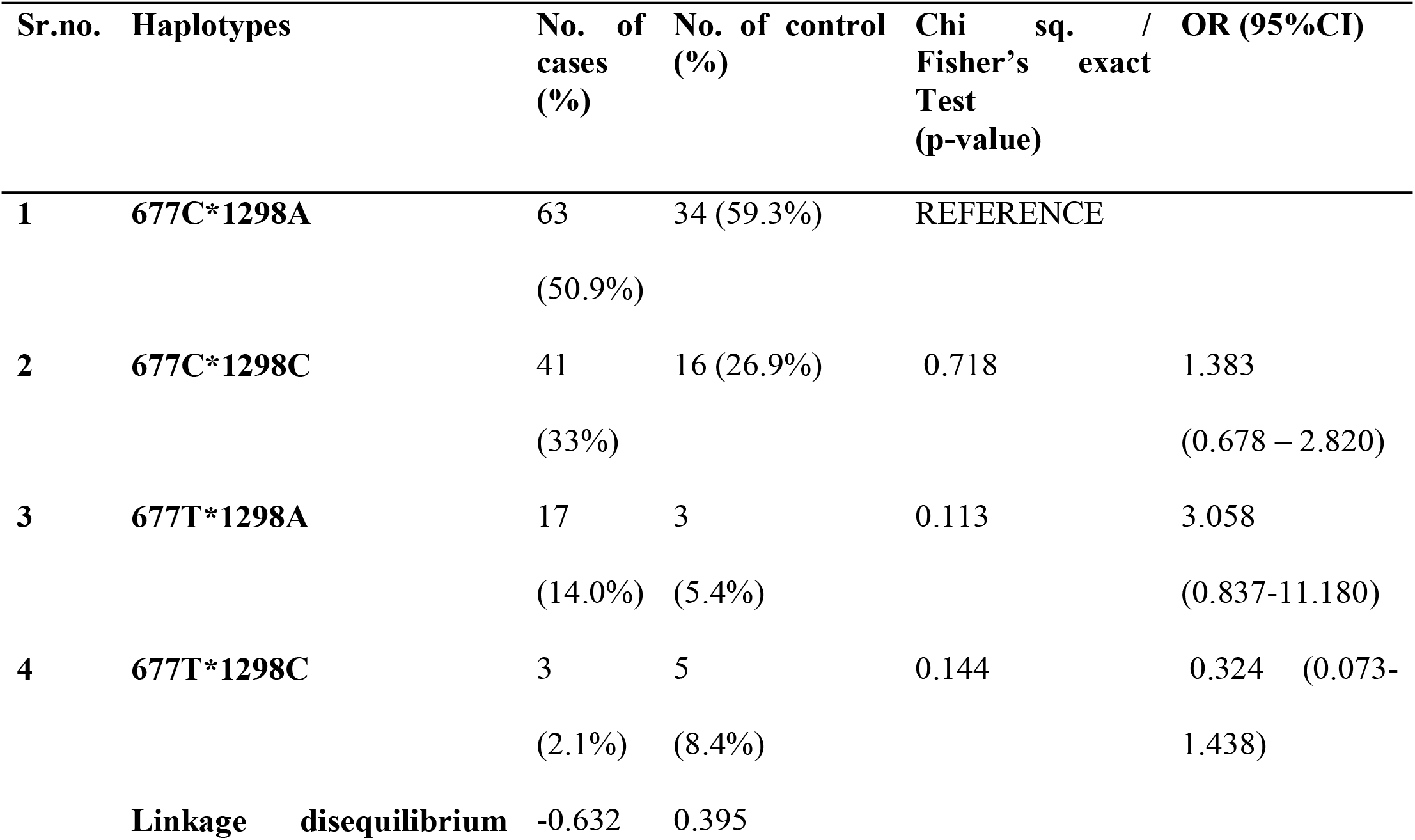
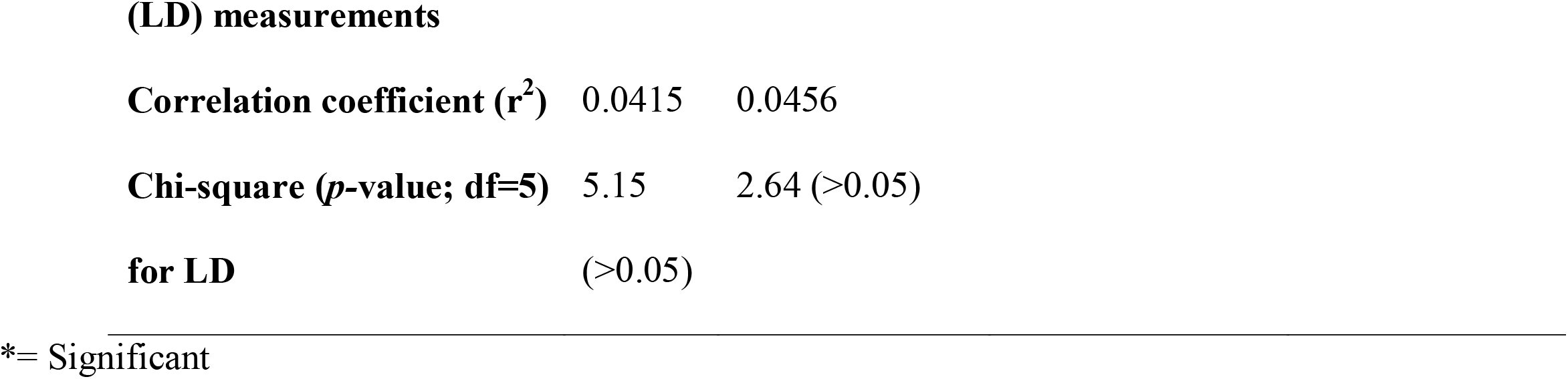
Semiparametric haplotype analysis-based and frequencies of *MTHFR* 677CT and 1298AC haplotypes in breast cancer patients and control.

## DISCUSSION

Breast cancers are a heterogeneous group of disorders, which are characterized by abnormal cellular proliferation in the mammary tissue [27]. The contribution/s of genetic factors are complex in nature, which can vary in penetrance [28]. The present study is a component of the breast cancer molecular genetic investigation study in Pakistani population. The aim is to decipher the molecular and genetic architecture of the breast cancers in a population, characterized by high consanguinity, multiparity, and low exposure to putative risk factors for breast cancers, and particularly low levels of alcohol and pork consumption. Thus, the present population provides a unique model for the epidemiological and molecular investigations of breast cancers. Additionally, folate deficiency is an established severe public health concern in women of reproducible ages in the studied population [29]. This insufficiency, often attributed to low intake, is likely to be compounded by the genetic factors, with consequent effect on the breast cancer spectrum observed across Pakistani population.

In the present study, we report lack of independent significant associations of the *MTHFR* 677C/T (rs 1801133) and 1298A/C (rs 1801131) polymorphisms with the risk of breast cancers in Pakistani population. However, to the best of our knowledge, two diplotype combinations show unique associations with breast cancer susceptibility in this population, which have not been reported earlier. The diplotype analysis revealed that 677CC+1298AC increased the risk of breast cancers significantly [OR: 2.553 (95% CI: 1.177-5.541)], while 677CT+1298AA had a significant protective effect [OR: 0.537 (95% CI: 0.404-0.713)].

The other important observation in the present study is the lack of *MTHFR* 1298CC genotype in breast cancer patients.

Three genotype combinations are absent in the studied breast cancer patients: CT/CC, CC/CC, and TT/CC. Interestingly, TT/CC combination was absent in controls and RCC patients as well [22].

The linkage equilibrium between two loci, which are in close proximity is a unique finding of the current report. In 1000 genome project [30], among South-Asian populations, these two SNPS have been shown to be in LD in PJL (Punjabis from Lahore), GIH (Gujrati Indians in Houston), ITU (Indian Telugu from the UK) populations; whereas these are known to be not associated in BEB (Bengali from Bangladesh), and STU (SriLankan Tamil from the UK). In the present study, in contrast to PJL, where D’ has been shown to be 1, and r^2^ to be 0.119, for breast cancer patients these values were -0.502, and 0.0273, respectively. In case of controls, the values were 0.41 and 0.0496, respectively. Thereby, lack of association between the variants at studied loci is reported in the present study. Thus, underscoring the importance of validating data from genome wide association studies.

## Conclusions

The present study reports unique associations of the combined *MTHFR* 677C/T and *MTHFR* 1298A/C SNPs with breast cancers in Pakistani population. A distinctive lack of association over the short genomic sequence is also a significant finding of the present molecular investigative report. The results have important implications in devising folate pathway specific strategies for breast cancer management in population sub-groups.

## Supporting information

STROBE CHECKLIST

## Data Availability

Data available upon request

## Acknowledgements

The authors wish to thank Pakistan Health Research Council for partial funding of the study. ICCBS for core facilities. AEMC, JPMC staff for their co-operation. The authors are especially grateful to the participants in the study.

## Declarations

### Data Availability

All data is available on request.

### Code Availability

Not Applicable.

### Authors’ Contribution

SA: Conceptualization, Sampling, Benchwork, Medical Record Data Collection, Analysis, Manuscript Writing Final Draft, Sponsorship.

SZZ: Case Sampling, Benchwork, Medical Record Data Collection, Analysis, Manuscript Writing Initial Draft.

SMA: Case Sampling, Benchwork, Medical Record Data Collection.

AS: Case Sampling, and Medical Record Data Collection.

MAM: Case Sampling, and Medical Record Data Collection.

AA, SK: Control Sampling, Benchwork.

### Ethical Approval

The study was conducted in accordance with the Declaration of Helsinki [20]. The project was approved by the ethical review committees (ERCs) of the participating institutions: the independent ERC, International Center for Chemical and Biological Sciences (ICCBS), University of Karachi, Karachi, Pakistan [ICCBS/IEC-016-BS/HT-2016/Protocol/1.0], and the Atomic Energy Medical Centre (AEMC), Jinnah Postgraduate Medical Centre (JPMC), Karachi, Pakistan [Admin-3(257)/2016]. All the samples were collected after obtaining written informed consent from each participant. The ethical approval from controls has been submitted and published earlier.

### Informed Consent

Informed consent was volunteered by all the participants on signed consent forms.

### Authors’ Approval

All the contributing authors approved the final draft.

